# Prevalence of educational needs and factors associated with educational severity among Rohingya refugee children in Bangladesh: a representative cross-sectional household survey

**DOI:** 10.64898/2026.01.12.26343959

**Authors:** Harry J Wilson, Nyanhial Yang, Md Nazmul Karim

## Abstract

**Background:** Approximately 1.2 million Rohingya refugees currently reside among the 33 refugee camps in Cox’s Bazar - Bangladesh, where the right to education remains a critical humanitarian concern. This study aimed to estimate the prevalence of Rohingya refugee children in educational need, elucidate predominant reasons among children in need, and identify factors associated with educational severity to inform targeted humanitarian relief efforts.

**Methods:** This study utilized participant data collected during the 2023 Joint Multi-Sectoral Needs Assessment—a representative cross-sectional household survey. Education-age children were assigned a severity score categorized as stressed, severe, or extreme based on proxy responses capturing educational access, learning conditions, and protection factors. Ordinal logistic regression examined factors associated with increasing educational severity scores.

**Findings:** Among 8,408 education age children surveyed from 2,959 households, 29.6% were in severe educational need and 10.1% were in extreme educational need. After adjustment, increasing educational severity scores were significantly associated with head of household age 45-59 (aOR= 1.29 [95%CI: 1.09–1.54]), head of household age 60+ (aOR= 1.31 [95%CI: 1.07–1.60]), female headed households (aOR= 1.33 [95%CI: 1.15–1.54]), early education age boys (aOR= 3.95 [95%CI: 3.21–4.85]) and teenage boys (aOR= 4.42 [95%CI: 3.73–5.24]) relative to primary school age boys, early education age girls (aOR= 4.23 [95%CI: 3.36–5.31]) and teenage girls (aOR= 19.06 [95%CI: 15.70–23.15]) relative to primary school age girls, primary school age girls (aOR= 0.82 [95%CI: 0.68–0.99]) relative to primary school age boys, and teenage girls (aOR= 3.54 [95%CI: 3.04–4.12]) relative to teenage boys.

**Interpretation:** Educational needs remain highly prevalent among Rohingya refugee children influenced by structural factors, systemic challenges in service provision, household vulnerabilities and serious child protection issues – particularly among teenage girls. Local humanitarian authorities are currently providing targeted relief to those in educational need but these interventions are threatened by sweeping international funding cuts.

**Research in Context:** *Evidence before this study:* Approximately half of the 1.2 million Rohingya refugees in Cox’s Bazar are education-age children. Contemporary humanitarian frameworks conceptualise education-in-emergencies as both a critical service and protective system to safeguard vulnerable children. Concerningly, prior analysis identified the education sector as the leading reason for households in extreme humanitarian need. Preliminary investigation foreshadowing gendered disparities and raised serious child protection concerns. However, these results were restricted to extreme needs only and did not disaggregate educational severity by age, gender, or underlying reason—limiting its ability to strategically inform sectoral programming.

*Added value of this study:* Sub-group analysis revealed nuanced age-gender dynamics with similarities observed among early education (age 3-4) and primary school (age 5-11) children, followed by stark disparities during adolescence between teenage boys and girls (age 12-18). Teenage girls experienced dramatically elevated odds of educational need, predominantly attributed to deprioritisation, inadequate gender separation, and serious protection concerns including child marriage and early pregnancy. Logistic regression quantified age-gender characteristics associated with educational severity whilst controlling for household vulnerability factors.

*Implications of all the available evidence:* Our findings demonstrate high prevalence of educational needs and pronounced gender inequities concentrated among teenage girls. The evidence suggests that humanitarian programming should be consolidated for younger children (age 3-11) and differentiated for teenagers (age 12-18) to address disparate needs through gender-specific interventions. The comparative strength of individual characteristics associated with educational severity suggests prevailing structural factors and systemic service delivery challenges, consistent with prior evidence of limited gender-responsive learning opportunities. These challenges are further exacerbated by large-scale facility closures that local humanitarian authorities attribute to sweeping international funding cuts. Strategic, intensive, and sustained relief efforts are needed, alongside longitudinal studies to monitor trends and clarify causal pathways.

## Introduction

Eight years on from the mass displacement events in late 2017, approximately 1.2 million Rohingya refugees are currently sheltering within 33 congested camps of Cox’s Bazar near the Bangladesh-Myanmar border.^1^ The protracted nature of the Rohingya refugee crisis underscores the necessary operational transition from an acute emergency response to an enduring humanitarian program. Meeting the humanitarian needs of Rohingya refugees remains imperative for the international community but has met a critical juncture characterised by entrenched vulnerabilities, aid dependencies, concurrent domestic needs to support host communities, deteriorating humanitarian conditions within the camps, and a resurgence in targeted violence across the broader in Myanmar that has displaced an additional 150,000 Rohingya.^2-6^ Although the contemporary humanitarian scenario has worsened, the international attention to the protracted crisis has waned.^2,4,6^ The timing of these operational challenges precipitate amid a backdrop of changing geopolitical landscapes and receding humanitarian budgets that continues to threaten the delivery of essential humanitarian relief services for Rohingya refugees.^3,5,7-9^

Approximately half of all Rohingya refugees sheltering among the camps in Cox’s Bazar are education age children. Consequentially, the demographic profile positions the education sector as a key cornerstone of the contemporary humanitarian response.^5,9^ Beyond the direct individual benefits, ensuring the right to education is of paramount importance to protect vulnerable children, foster community resilience, build capacity for self-determination and strengthen prospective recovery efforts when circumstances allow safe voluntary repatriation.^5,6^ A growing body of evidence highlights the pragmatic need for standardized education in emergency programs to incorporate nuanced age-gender responsive interventions that are co-designed with refugee communities, contextually framed within the lived experience of education age children they serve, and commensurate with the specific barriers they endure.^10-14^ Additionally, the transition from childhood to adolescence intensify gendered effects among school-age boys and girls that may differently interact with standardized education services. This developmental period is particularly crucial for teenage girls, significantly influencing their physical, psychosocial, and emotional well-being that persist into adulthood.^14^ Reciprocally, the implications of compromised educational attainment are compounded in protracted displacement settings such as the Rohingya refugee crisis where cumulative exposure over time threatens to not only widen but entrench gendered inequities.^14,15^ Concerningly, prior analysis has identified the education sector as the leading reason for extreme humanitarian needs among Rohingya refugee households surveyed during the 2023 Joint Multi-Sectoral Needs Assessment (J-MSNA).^16^ Preliminary investigation foreshadowed gendered disparities and raised serious child protection concerns.^16^ However, this analysis was restricted to extreme educational needs only and did not disaggregate education sector severity scores by age, gender or underlying reasons that limits its ability to strategically inform sectoral programming.^16^

Appling a more sensitive sector-specific investigation of the education sector using the same 2023 J-MSNA dataset, this study aimed to estimate the prevalence of Rohingya refugee children in educational need, evaluate the distribution of education sector severity by age, gender and attributable reason to intelligently inform specific interventions, and identify household vulnerability factors and demographic characteristics associated with educational severity scores to strengthen targeted relief efforts by local humanitarian authorities.^17^

## Methods

### Study design, participants and sampling strategy

This study draws upon anonymized participant data from the 2023 J-MSNA, a representative cross-sectional survey of Rohingya households within the 33 refugee camps of Cox’s Bazar, used for secondary analysis.^17^ The primary data collection was conducted by the REACH Initiative in conjunction with sector-specific authorities and local enumerators. The dataset was accessed on the 27th of March 2025 under license approved by UNHCR. The study population included all Rohingya refugee households registered within the 33 camps of Cox’s Bazar.^18,19^ An administratively complete sampling frame of registered households was compiled by camp authorities.^18,19^ A total of 3,465 households were selected for data collection using stratified random sampling with camps as strata ^18,19^ The number of households selected for sampling at each camp were guided by a single proportion sample size calculation that derived the minimum number of households needed to ensure camp-specific results could be concluded with a minimum level of precision (10% margin of error) at a 95% confidence level.^18,19^ The sample size was adjusted for the finite household population that were eligible for selection at each camp and inflated by an additional 10% to mitigate non-response.^18,19^ Although the study was primarily powered to enable camp specific results at the household-level, the achieved sample size provided sufficient precision for individual-level subgroup analyses by age and gender. Of the 3,465 households selected for sampling, a total sample size of 3,400 households were surveyed between August 27th and September 17th 2023 with individual data pertaining to 18,172 household members. Missingness was minimal (<1%) and these records were excluded from analysis.

### Data collection

Data were collected through in-person household interviews administered by trained enumerators using a validated structured questionnaire that captured standardized humanitarian indicators and sector-specific items.^18,19^ Gender-balanced field teams (comprising male and female interviewers) conducted interviews with an adult (age 18 or older) respondent of the same gender (self-identified).^18,19^. Education status and reported reasons for educational need were provided by the adult respondent on behalf of each child within the household (proxy respondent), rather than by the children themselves. Responses were digitally recorded using the KoBo Collect application on mobile devices and routinely uploaded to a secure UNHCR server.^18,19^ Raw participant data was cleaned according to the REACH Initiative’s minimum standards protocol with a final de-identified dataset provided for statistical analysis.^18,19^ Verbal informed consent was secured from all household respondents and formally recorded in the deidentified dataset. Respondents were informed of the survey’s objectives, the voluntary nature of their participation, measures to protect response confidentiality and prerogative to decline any question or discontinue the interview at any time. Enumerators were educated on the principles of participant safeguarding and trained to provide formal referral protocols upon discovery of urgent protection or health concerns.

### Variables

#### Outcome variables

The primary outcome was education severity score assigned to each education age child (age 3-18) based on responses to questionnaire items that capture the four dimensions of educational access, learning conditions, protection and aggravating circumstances. Education severity scores were assigned based on recommended criteria outlined in the Global Education Cluster (UNICEF and Save the Children) severity classification manual, Joint Intersectoral Analysis Framework (JIAF) and Multi-Sectoral Needs Index (MSNI).^18,20,21^ Severity was scored according to the JIAF five-point intersectoral severity classification scale ranging from minimal (score=1) to catastrophic (score=5). The five-point intersectoral severity scale was operationalised as a three-point scale in line with J-MSNA methods due to contextual factors and sampling units rather than inferential constraints that may otherwise compromise the construct validity of the established classification scheme. The minimal severity category (score=1) could not be assigned due to the absence of formal education services within the camps. Hence, the lowest possible score that could be assigned reflected regular attendance at a non-formal learning centre corresponding to a severity score of stressed (score=2) rather than minimal (score=1). Conversely, the catastrophic severity score (score=5) requires aggregate response-level data indicating complete sectoral collapse and systemic exclusion rather than individual learning conditions and reasons for not accessing education as reported by proxy respondents on behalf of the household. The operationalized three-point scale categorised each education age child as either stressed (score=2), severe (score=3) or extreme (score=4). The restricted three-point scale retains the same ordinal severity distinctions of the established five-point intersectoral severity scale, supporting meaningful comparisons of educational severity scores across subgroups. The lowest possible score of stressed (score=2) was defined as accessing non-formal education in a protected environment offering acceptable learning conditions. Severe (score=3) was classified as either not accessing education due to non-aggravating circumstances, or accessing non-formal education in a protected environment but experiencing poor learning conditions or minor safety issues that do not present an immediate threat to the child’s wellbeing. Extreme (score=4) was defined as either not accessing education services due to aggravating circumstances, or attending an unprotected learning environment due to serious safety issues that endanger the child’s well-being. Aggravating circumstances correspond to the most severe accessibility barriers such as marriage or pregnancy, child paid and unpaid labour to support the household instead of attending school, recruitment by armed groups, discrimination, and safety threats whilst travelling to or attending school.^18,21^ Unprotected learning environment included armed group recruitment, school attacks, gender based or sexual violence, and discrimination based on the child’s ethnic, gender or religious identity.^20,21^ The underlying reason for each child’s education severity score were utilised as secondary outcomes to evaluate predominate underlying reasons for educational severity scores and their distribution across age-gender subgroups.

#### Independent variables

Individual characteristics of education age children and household vulnerability factors evaluated for association with educational needs were identified from previous J-MSNA research, Refugee Influx Emergency Vulnerability Assessments (REVA), and funding impact analysis conducted by the World Food Programme.^7,8,22,23^ Age groups were assigned according to previous camp levels findings and appropriate cutoff points identified from trends observed over increasing age.^24^ Subgroups among education age children (age 3-18) included early education age children (age 3-4), primary school age children (age 5-11) and teenage children (age 12-18). Head of household age was categorized into 15-year age groups. Marital status was collapsed into a binary variable that included married and single participants (no partner/separated/divorced/widowed) due to limited observations. Head of household disability status was classified as a binary variable corresponding to participants citing either “a lot of difficulty” or “cannot do it at all” to any of the six functional domains as per the Washington Group Short Set (WG-SS) guidelines.^25^ Monthly household income per capita (household members) was categorized according to quartiles and utilized to investigate associations with financial vulnerability.

### Statistical analysis

All results were computed using Stata version 19.5 survey functions that accounted for the stratified sampling design, adjusted for the clustering of education age children within the same household and controlled for differential probability of household selection between the camps (strata) using survey weights computed as the inverse probability of selection.^26^

Descriptive statistics were used to analyse demographic characteristics of sampled households and education age children. Weighted frequencies and percentages were reported for categorical variables. Median and interquartile range (IQR) or mean and standard deviation (SD) were reported for continuous variables depending on symmetry. Prevalence estimates of children and households experiencing educational needs were reported as weighted percentages with 95% confidence intervals (95%CIs) for precision.

Education severity scores and their attributable reasons were disaggregated by age and gender. The distribution of severity scores and reported reasons were assessed over increasing age and by gender to assess linear trends. Ordinal logistic regression analysis was conducted to investigate factors associated with increasing education sector severity scores and the assumption of proportional odds was tested. Unadjusted odds ratios (OR) and adjusted odds ratios (aOR) were reported for bivariate and multivariate estimates respectively. A threshold value of p ≤ 0.05 was used to conclude statistical significance and 95%CIs were reported for precision using design-adjusted standard errors computed via Taylor series linearization.

Potential effect modification was considered between head of household gender and marital status to investigate concentrated effects among single female headed households. Two-way age and gender interactions among education age children (age-gender and gender-age) were investigated using both stratified analysis and interaction terms. Gender-age interactions were investigated using stratified (by gender) analysis reporting the relative risk associated with age groups among boys and among girls. Age-gender interactions were investigated using interaction terms to evaluate the relative risk associated with girls relative to boys within the same age group. Pairwise correlation and variance inflation factor (VIF) analysis were conducted to evaluate potential problematic multicollinearity.

## Results

### Participant characteristics

Among the 3,400 total households sampled, 441 households did not contain education age children (age 3-18) and were excluded from subsequent analysis (Table 1). A total of 8,408 education age children were surveyed from the remaining 2,959 households across the 33 camps. The number of households without education age children (n=441) did not significantly differ between the camps (p >0.05). The proportional contribution of each camp to the overall sample showed minimal variation (mean change= +0.5%; range: -12.6% to +8.7%) before (N=3,400) and after (N=2,959) the exclusion of households without education age children. The number of households and education age children sampled per camp was approximately normally distributed with a mean of 90 households (SD= 35; Table 1) and 255 education age children (SD= 95; Table 1) respectively. The median head of household age was 39 years (IQR: 30 **–** 50; Table 1) and the majority of households were male headed (79.8%; Table 1). Approximately half of surveyed children were aged between 5-11 years (47.4%; Table 1) and the proportion of boys sampled (50.3%; Table 1) was slightly higher than the proportion of girls (49.7%; Table 1). Less than two-thirds of education age children were enrolled and regularly attending informal education within the camps (60.3%; Table 1). The prevalence of children in educational need was 39.7% (95% CI: 38.4% – 41.0%) and almost two-thirds of households contained at least one child in educational need (65.7% [95% CI: 63.8% – 67.5%]).

**Table 1:** Characteristics of sampled households (N= 2,959) and education age children (N= 8,408)

|  | Summary* |
| --- | --- |
| **Households per Camp, Mean (N= 2,959)** | 90 (SD: 35) |
| **Education Age Children per Camp, Mean (N= 8,408)** | 255 (SD: 95) |
| **Head of Household Age, Median (N= 2,959)** | 39 [IQR: 30 – 50] |
| 18-29 | 688 (23.2%) |
| 30-49 | 1414 (47.8%) |
| 50+ | 857 (29.0%) |
| **Head of Household Gender (N= 2,959)** |  |
| Male | 2361 (79.8%) |
| Female | 598 (20.2%) |
| **Education Age Children per Household, Median (N= 2,959)** | 3 [IQR: 2 – 4] |
| 1-2 | 1465 (49.5%) |
| 3-5 | 1284 (43.4%) |
| 6-9 | 210 (7.1%) |
| **Age, median (N= 8,408)** | 10 [IQR: 6 – 14] |
| 3-4 | 1,165 (13.9%) |
| 5-11 | 3,985 (47.4%) |
| 12-18 | 3,258 (38.7%) |
| **Gender (N= 8,408)** |  |
| Boys | 4233 (50.3%) |
| Girls | 4175 (49.7%) |
| **Education Sector Severity (N= 8,408)** |  |
| Stressed (Severity Score=2) | 5070 (60.3%) |
| Severe (Severity Score=3) | 2488 (29.6%) |
| Extreme (Severity Score=4) | 850 (10.1%) |
| **Prevalence of Educational Need <sup>a</sup>** |  |
| Children in Educational Need (N= 8,408) | 39.7% [95% CI: 38.4% – 41.0%] |
| Households in Educational Need (N= 2,959) | 65.7% [95% CI: 63.8% – 67.5%] |

\* Estimates are relative to either the total number of households with education age children (N= 2,959) or the total number of education age children (N= 8,408). Summary statistics are weighted frequency and weighted percentages for categorical variables, weighted median (IQR) for asymmetrically distributed continuous variables, weighted mean (SD) for symmetrical continuous variables, or weighted population prevalence (%) and 95% CIs for outcome variables.
<sup>a</sup> Children in educational need was classified as severe or extreme education severity scores (severity score=3+) as per MSNI methods.<sup>18</sup> Children in education need defined as per JIAF and education cluster guidelines - Education age children in the areas affected by crisis who do not have access to protective education and acceptable learning conditions, which can negatively impact (i) their physical and psychosocial wellbeing, (ii) cognitive development, and (iii) their ability to meet their future needs.<sup>20,21</sup> Households in educational need categorised as those with at least one child in educational need as per MSNI methods.<sup>18</sup>

**Table 2:** Individual factors and household characteristics associated with higher educational severity scores.

|  | Summary | Odds Associated with Higher Educational Severity Scores |  |  |  |
| --- | --- | --- | --- | --- | --- |
|  | (N=8,408) | Unadjusted |  | Adjusted |  |
|  | n (%) | OR (95%CI) | p-value | aOR (95%CI) | p-value |
| **Head of Household Age** |  |  |  |  |  |
| 18-34 | 2,521 (30.0%) | 1.00 | – | 1.00 | – |
| 35-44 | 2,577 (30.6%) | **1.28 (1.10–1.47)** | **<0.001\*** | 0.92 (0.77–1.09) | 0.319 |
| 45-59 | 2,225 (26.5%) | **2.39 (2.08–2.75)** | **<0.001\*** | **1.29 (1.09–1.54)** | **0.003\*** |
| 60+ | 1,085 (12.9%) | **2.80 (2.36–3.31)** | **<0.001\*** | **1.31 (1.07–1.60)** | **0.008\*** |
| **Head of Household Gender** |  |  |  |  |  |
| Male | 6,925 (82.4%) | 1.00 | – | 1.00 | – |
| Female | 1,483 (17.6%) | **1.50 (1.31–1.72)** | **<0.001\*** | **1.33 (1.15–1.54)** | **<0.001\*** |
| Female: Married <sup>a</sup> | 780 (9.3%) | 1.00 | – | 1.00 | – |
| Female: Single/Divorced/Widowed <sup>a</sup> | 703 (8.3%) | 1.15 (0.91–1.46) | 0.241 | 1.01 (0.78–1.30) | 0.960 |
| **Head of Household Disability Status** |  |  |  |  |  |
| No Disability | 6,972 (82.9%) | 1.00 | – | 1.00 | – |
| Living with a Disability | 1,436 (17.1%) | **1.19 (1.03–1.37)** | **0.017\*** | 1.03 (0.88–1.21) | 0.686 |
| **Monthly Household Income (per capita)** |  |  |  |  |  |
| 1<sup>st</sup> Quartile | 2,385 (28.4%) | 1.00 | – | 1.00 | – |
| 2<sup>nd</sup> Quartile | 2,379 (28.3%) | 0.99 (0.86–1.14) | 0.903 | 1.09 (0.93–1.28) | 0.280 |
| 3<sup>rd</sup> Quartile | 2,174 (25.9%) | 0.90 (0.77–1.04) | 0.148 | 0.95 (0.81–1.12) | 0.543 |
| 4<sup>th</sup> Quartile | 1,470 (17.4%) | 1.08 (0.92–1.26) | 0.363 | 1.13 (0.95–1.35) | 0.157 |
| **Boys: Age Group** |  |  |  |  |  |
| Boys: Age 5-11 | 2,053 (24.4%) | 1.00 | – | 1.00 | – |
| Boys: Age 3-4 | 579 (6.9%) | **3.84 (3.14–4.70)** | **<0.001\*** | **3.95 (3.21–4.85)** | **<0.001\*** |
| Boys: Age 12-18 | 1,601 (19%) | **4.82 (4.09–5.68)** | **<0.001\*** | **4.42 (3.73–5.24)** | **<0.001\*** |
| **Girls: Age Group** |  |  |  |  |  |
| Girls: Age 5-11 | 1,933 (23%) | 1.00 | – | 1.00 | – |
| Girls: Age: 3-4 | 585 (7%) | **4.12 (3.30–5.14)** | **<0.001\*** | **4.23 (3.36–5.31)** | **<0.001\*** |
| Girls: Age 12-18 | 1,657 (19.7%) | **21.23 (17.56–25.67)** | **<0.001\*** | **19.06 (15.70–23.15)** | **<0.001\*** |
| **Age 3-4: Gender** |  |  |  |  |  |
| Age 3-4: Boys | 579 (6.9%) | 1.00 | – | 1.00 | – |
| Age 3-4: Girls | 585 (7%) | 0.87 (0.70–1.09) | 0.221 | 0.88 (0.70–1.10) | 0.263 |
| **Age 5-11: Gender** |  |  |  |  |  |
| Age 5-11: Boys | 2,053 (24.4%) | 1.00 | – | 1.00 | – |
| Age 5-11: Girls | 1,933 (23%) | **0.81 (0.67–0.98)** | **0.033\*** | **0.82 (0.68–0.99)** | **0.044\*** |
| **Age 12-18: Gender** |  |  |  |  |  |
| Age 12-18: Boys | 1,601 (19%) | 1.00 | – | 1.00 | – |
| Age 12-18: Girls | 1,657 (19.7%) | **3.58 (3.07–4.17)** | **<0.001\*** | **3.54 (3.04–4.12)** | **<0.001\*** |

\* Statistically significant at $p \leq 0.05$ .<sup>a</sup> Additional estimates from a duplicate model with interaction terms applied to investigate effect modification among single/divorced/widowed female headed households relative to married female headed households. Note that estimates reported for female headed households represent the main effects among all female headed households rather than the subgroup of married female headed households. All multivariable estimates were adjusted for main effects only.

### Educational Severity by age and gender

The distribution of education sector severity scores (Fig. 1A) and their attributable reasons (Fig. 1B) disaggregated by age-group and gender demonstrated three clear patterns: (i) substantial unmet educational need in early childhood driven by enrolment barriers, (ii) high attendance among children aged 5–11 with limited extreme need, and (iii) a sharp decline in enrolment after age 12 with pronounced gender disparities. Approximately half (54.3%; Fig. 1A) of early education age children (3-4 years) were enrolled and regularly attending informal education with similar results observed among boys (52.4%) and girls (56.1%). An additional 40.4% were found to be in severe educational need mostly attributable to “unable to register or enrol” (26.5%; Fig. 1B), “too young” (5.3%) and “education is not a priority” (3.3%). A further 5.3% (Fig. 1A) of early education age children were experiencing extreme educational need mostly attributable to “protection risks while commuting to school” (4.3%; Fig. 1B). The majority of primary school age children (5-11) were enrolled and regularly attending informal education (83.3%; Fig. 1A) with similar results observed between boys (81.8%) and girls (84.9%). An estimated 13.9% of primary school age children (5-11) were in severe educational need predominantly attributable to “attending madrasa instead” (4.9%; Fig. 1B), especially among boys (7.4%) compared to girls (2.3%). A relatively small proportion of primary school age children (5-11) were found to be in extreme educational need (2.8%; Fig. 1A). In contrast, only a third (34.4%; Fig. 1A) of teenage children age 12-18 were enrolled and regularly attending informal education with significant heterogeneity observed between boys (49.1%) and girls (20.1%). Most teenage children were experiencing severe educational needs (44.9%), with greater prevalence observed among teenage girls (50.6%) compared to boys (38.9%). Concerningly, a higher proportion of teenage girls were in extreme educational need (29.3%) than enrolled and regularly attending informal education (20.1%).

**Fig. 1A:**
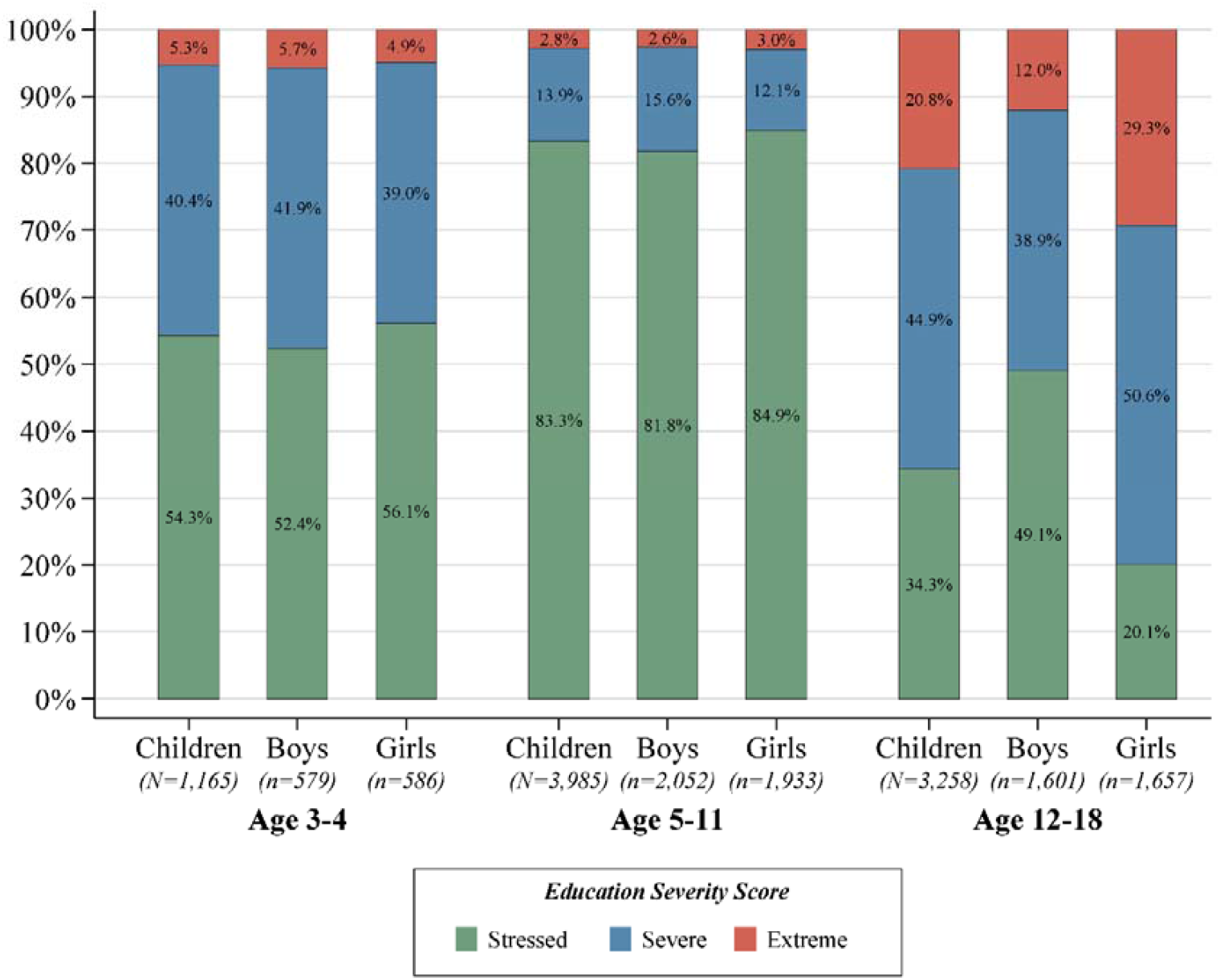
Education severity scores among education age children by age group and gender.

**Fig. 1B:**
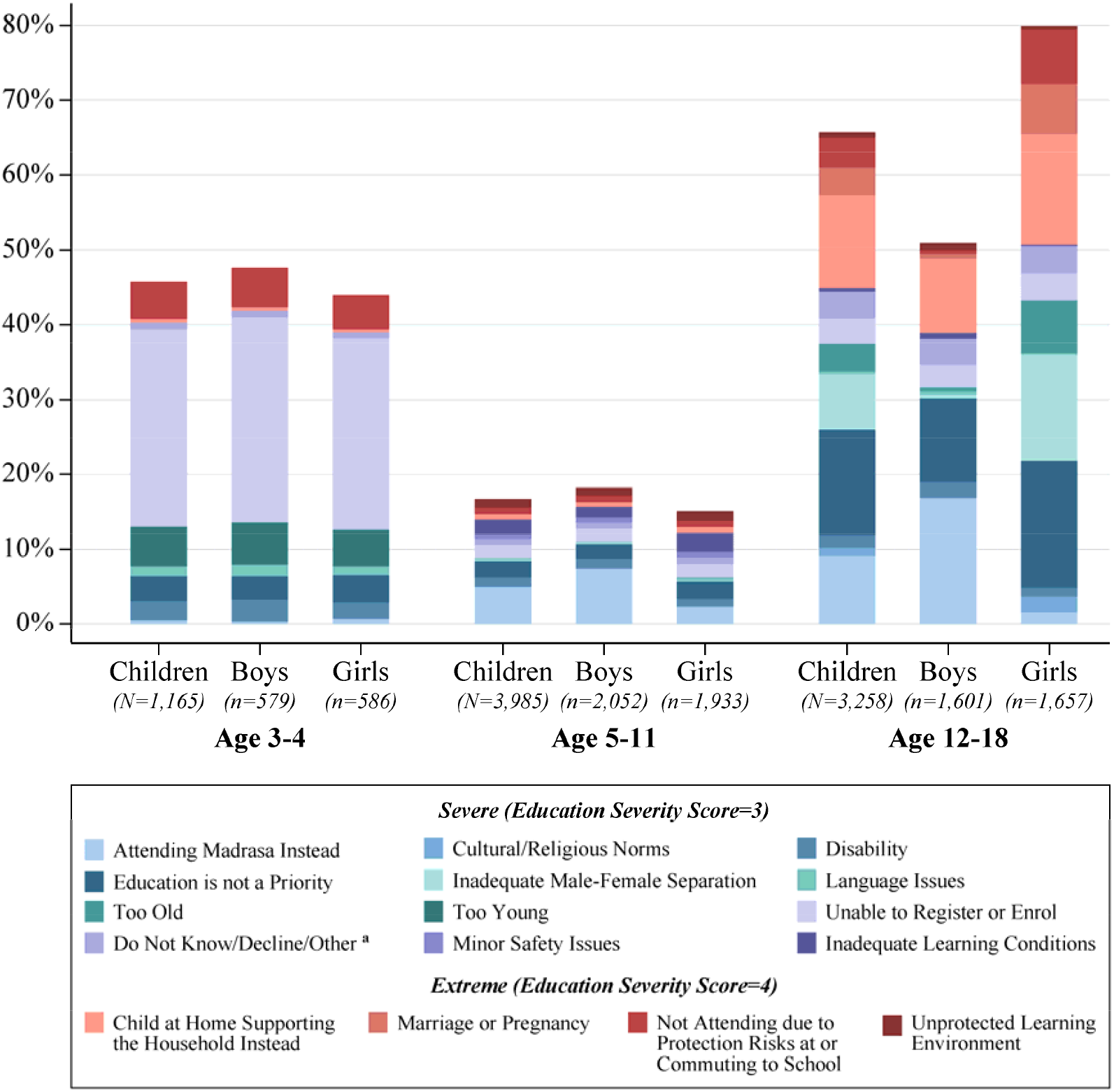
Attributable reason for educational needs among children (age 3-18) by age group and gender. ^**a**^ Rarely reported reasons (<1%) for severe educational needs among all cohorts were recoded as “other” rather than individually categorised for visual clarity. “Other” includes the following reasons and percentages: **Children Age 3-4:** Lack of Schools (0.3%), Language Issues (1.3%), Decline/Do not Know (0.6%) **Boys Age 3-4:** Lack of Schools (0.2%), Language Issues (1.5%), Decline/Do not Know (0.7%) **Girls Age 3-4:** Lack of Schools (0.4%), Language Issues (1.2%), Decline/Do not Know (0.4%) **Children Age 5-11:** Illness (0.1%), Lack of Schools (0.2%), Decline/Do not Know (0.5%) **Boys Age 5-11:** Cannot Afford Education Costs (0.1%), Illness (0.1%), Lack of Schools (0.1%), Decline/Do not Know (0.5%) **Girls Age 5-11:** Illness (0.1%), Lack of Schools (0.4%), Decline/Do not Know (0.4%) **Children Age 12-18:** Cannot Afford Education Costs (0.2%), Child Uninterested (0.3%), Home Based Learning (0.4%), Inadequate Curriculum (0.2%), Decline/Do not Know (2.4%) **Boys Age 12-18:** Cannot Afford Education Costs (0.3%), Child Uninterested (0.5%), Home Based Learning (0.7%), Inadequate Curriculum (0.1%), Decline/Do not Know (1.8%) **Girls Age 12-18:** Cannot Afford Education Costs (0.1%), Child Uninterested (0.1%), Home Based Learning (0.1%), Illness (0.1%), Inadequate Curriculum (0.2%), Decline/Do not Know (3.0%)

The analysis of education severity scores over increasing age and by gender illustrated that boys and girls were similar between ages 3-11 (Fig. 2A). An inverse proportional relationship was also evident between stressed and severe education severity scores with a steady proportion of children age 3-11 experiencing extreme educational needs. A sharp decline in the proportion of children enrolled and regularly attending school was observed over ages 12-18. Gendered disparities regarding severe and extreme educational needs emerged between ages 11-12 (Fig. 2A). A rapid rise in severe educational needs was observed among adolescent girls age 12-15 (Fig. 2A), predominantly attributable to “education is not a priority” and “inadequate male-female separation” as reported by household respondents (Fig. 2B). A linear trend between extreme educational needs and increasing age was evident among girls age 11-18 (Fig. 2A). The increasing trend in the proportion of girls age 11-15 experiencing extreme educational need was mostly attributable to “child at home supporting the household instead”, and not attending due to “protection risks at or commuting to school” (Fig. 2B). In contrast, the continuous rise in extreme educational needs among girls age 16-18 was primarily due to “marriage or pregnancy” (Fig. 2B). Educational needs among boys remained relatively constant between age 12-15 then increased between 16-18 predominantly attributable to “education is not a priority” (Fig. 2B) and “child at home supporting the household instead” (Fig. 2B).

**Fig. 2A:**
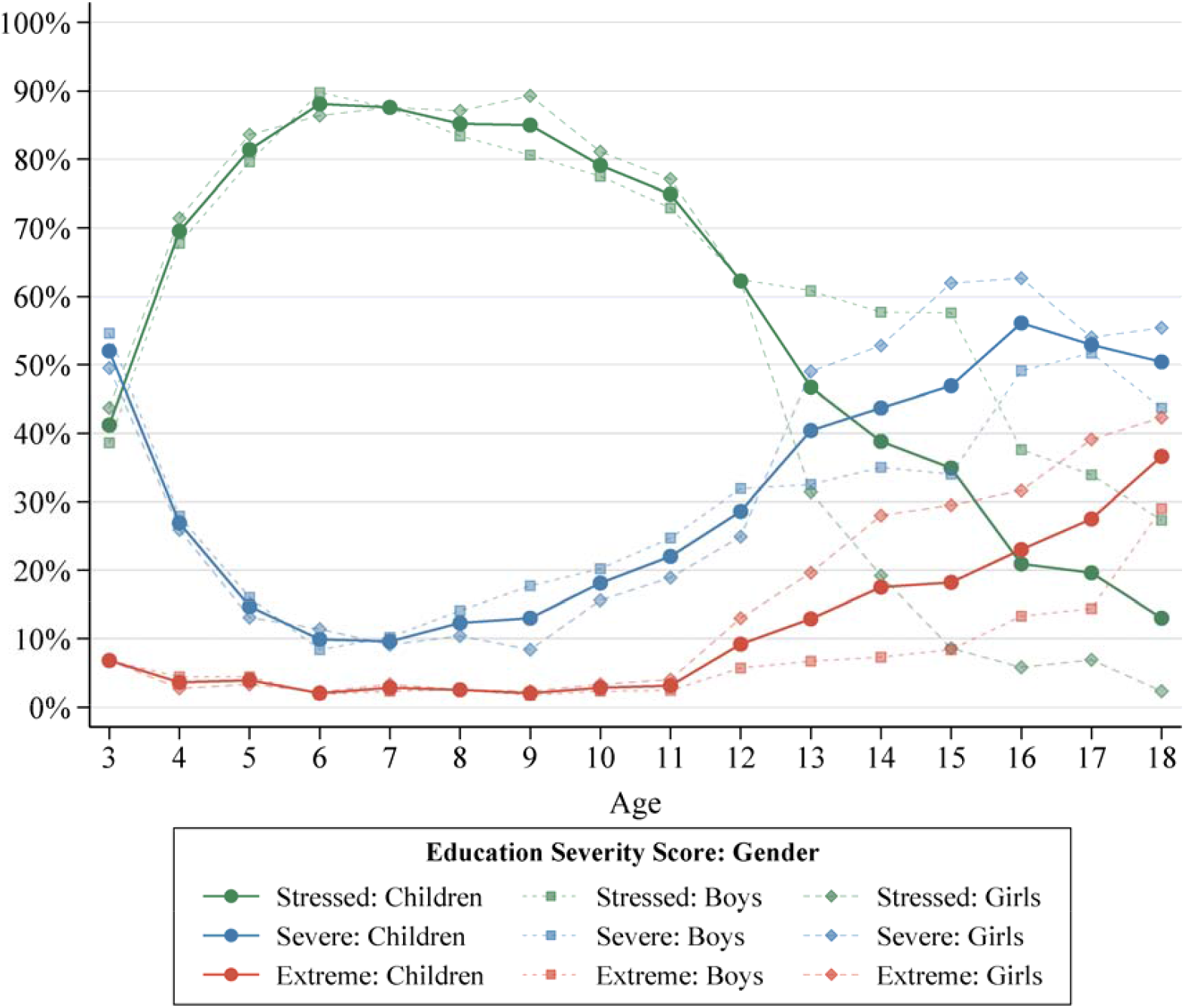
Education severity scores over increasing age (3-18) and by gender (children, boys, girls).

**Fig. 2B:**
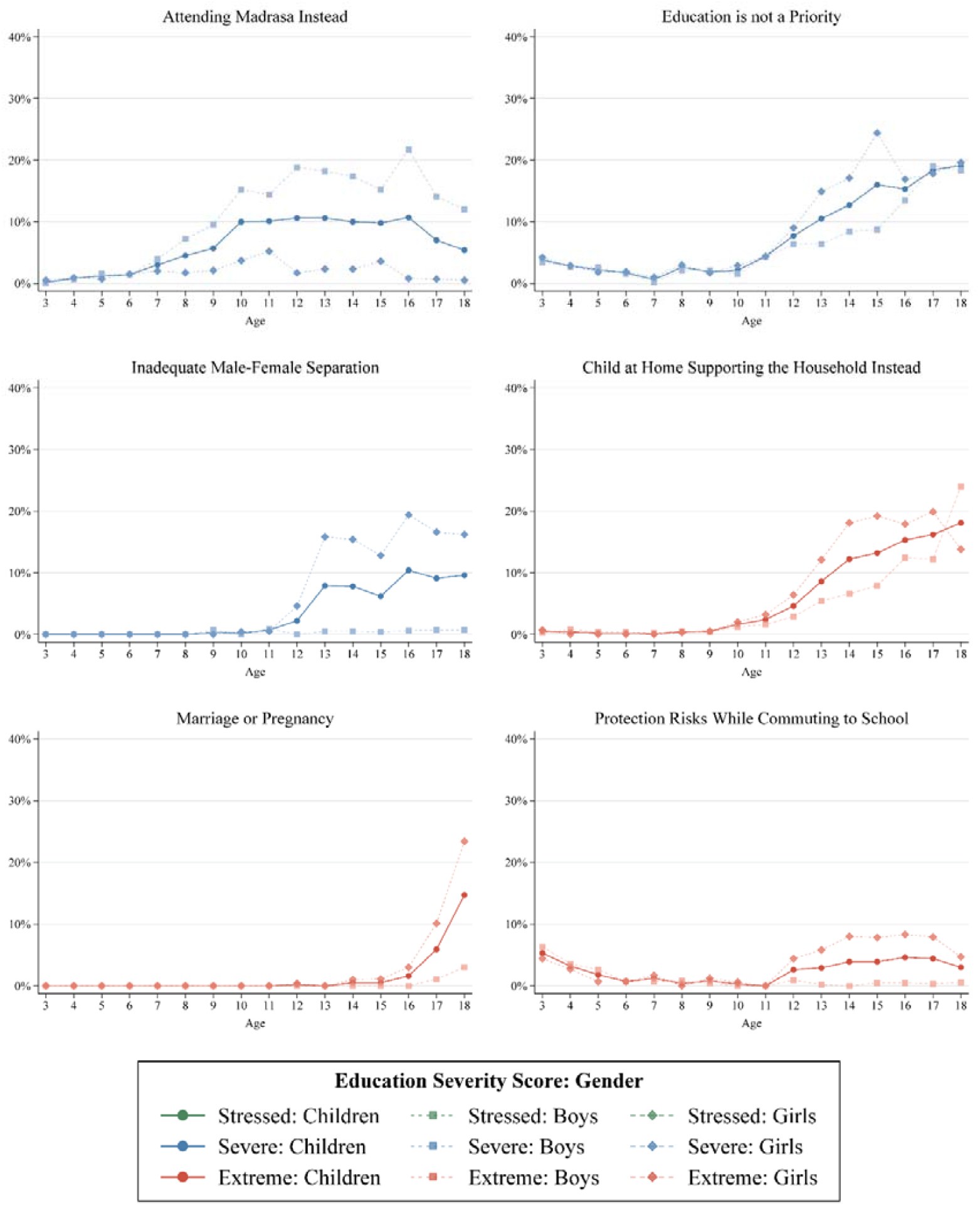
Reported reason for educational needs over increasing age (3-18) and by gender (children, boys, girls).

### Household vulnerability factors and individual characteristics associated with educational severity

In unadjusted analysis, increasing educational severity scores were associated with older head of households, female headed households, head of households living with a disability, age-groups among boys, age-groups among girls, and girls relative to boys within the same age group.

After adjustment in multivariable analysis, increasing educational severity scores were significantly associated with head of household age 45-59 (aOR= 1.29 [95%CI: 1.09–1.54]), head of household age 60+ (aOR= 1.31 [95%CI: 1.07–1.60]), female headed households (aOR= 1.33 [95%CI: 1.15–1.54]), early education age boys (aOR= 3.95 [95%CI: 3.21–4.85]) and teenage boys (aOR= 4.42 [95%CI: 3.73–5.24]) relative to primary school age boys, early education age girls (aOR= 4.23 [95%CI: 3.36– 5.31]) and teenage girls (aOR= 19.06 [95%CI: 15.70–23.15]) relative to primary school age girls, primary school age girls (aOR= 0.82 [95%CI: 0.68–0.99]) relative to primary school age boys, and teenage girls (aOR= 3.54 [95%CI: 3.04–4.12]) relative to teenage boys

The multivariate model indicated adequate fit over a null model (p<0.001) and did not violate the assumption of proportional odds. No evidence of problematic multicollinearity was observed, all pairwise correlations between independent variables were below 0.7 and the mean VIF was 1.5 with no extreme outliers (VIF_max_<10). No statistically significant evidence of an interaction on the multiplicative scale was found between female headed households and marital status (p >0.05) with main effects reported in consequence.

## Discussion

The demographic characteristics of households with education age children closely mirror those of all sampled households that prior analysis demonstrated to be strongly aligned with UNHCR’s database of all Rohingya refugees registered within the camps.^16^ The alignment observed between the sample and population characteristics strongly support the representativeness of participants and hence generalisability of our results.^16^

Despite ongoing humanitarian relief efforts by local authorities, the findings highlight that almost two-thirds of households, and 2-in-every-5 children age 3-18 were in educational need. Disaggregation by age group illustrated the higher prevalence of educational needs among early education age children (age 3-4) and teenage children (age 12-18). Further disaggregation by gender evidenced the similarities among boys and girls during childhood (age 3-11), followed by the rapid divergence of educational needs during adolescence (age 12-18). These age-gender dynamics are best illustrated by the distribution of severity scores and their attributable reasons over increasing age that further evidenced the gendered similarities during childhood and contrasting linear trends observed during adolescence that may reflect the emergence of gendered interactions that differentiate teenage boys and teenage girls. These descriptive interpretations are supported by the logistic regression results that revealed a sharp increase in the odds associated with teenage girls relative to both primary school age girls and teenage boys.

The associations observed between educational needs and individual age-gender characteristics persisted with limited attenuation after adjustment for common household vulnerability factors. The comparative strength of association with individual-level severity scores among education age children suggest the broader influence of exogenous structural factors and systemic challenges within the education sector. This interpretation is consistent with findings from previous sector-specific analysis that highlight the limited learning opportunities for teenage girls and lack of reintegration support for out-of-school teenagers transitioning from at-home to face-to-face learning as COVID-19 restrictions subsided.^5,10,11,15,22^ Similarly, our findings align with education-in-emergency frameworks that conceptualise schooling as both an important humanitarian service and protective system, with an emphasis on safeguarding the heightened vulnerability among adolescent children. In contrast, the elevated risk associated with female headed households, young adult (18-24) headed households and elderly (age 45+) headed households draw focus towards the survival pressures that may provoke households to deprioritise their children’s education. These interpretations are supported by the distribution of reported reasons for educational need, which are highly concentrated among teenage girls. The trends regarding deprioritization and deteriorating severity scores highlight the intersections between the education sector and the broader survival pressures within the camps that may position households to adopt negative livelihood coping strategies.^1,27^ In addition, the high proportion of teenage girls in extreme educational need reportedly due to childhood marriage and early pregnancy emphasize the close links between deteriorating educational needs and serious child protection issues that require urgent remediation efforts.

The findings should be interpreted within the context of multiple strengthens and limitations. The stratified random sampling, survey weighting and face-to-face interviews enhance representativeness of participants. However, the findings are predominantly limited by the use of self-reported data collected from proxy respondents on behalf of the household that may consequentially introduce measurement bias and undermine the accuracy of the results. The cross-sectional design limits causal inference and observed associations may reflect unmeasured confounding. These potential sources of confounding including several key determinants of educational access—such as learning-centre proximity, teacher availability, household mobility restrictions, and camp-level security conditions that were not directly measured. Finally, sensitive protection-related barriers (e.g., marriage, pregnancy, exploitation, safety threats) may be under-reported due to stigma and proxy reporting, potentially leading to conservative estimates of extreme need.

Our findings present numerous implications for humanitarian programming within the education sector. The age- and gender-specific gradients identified in this study suggest that education programming should be consolidated among early education and primary school age children, and differentiated among teenage boys and teenage girls to strategically target gendered protection issues, sociocultural barriers and accessibility constraints. In response, over 20 humanitarian projects coordinated by lead agencies, international authorities and non-government organisations are currently providing relief to Rohingya refugee children in educational need.^5^ These projects include school feeding programs for disadvantaged students, reuptake programs for out-of-school children, accelerated learning programs for students who are at risk of falling further behind and restructuring of learning centres to provide female-only classrooms that redress sociocultural barriers among teenage girls.^4,9^ The continuity of these projects are critical to affirm the right to education among Rohingya refugee children yet remain desperately underfunded at just 15% of their stated requirements.^28^ The detrimental impacts of funding cuts among the shock-prone education sector are already evident, provoking the closure of 43% of learning facilities and denying access to education for approximately 200,000 Rohingya children.^1,29^ Rapid restructuring efforts in conjunction with community adaptation have secured the gradual recovery of the education sector.^1^ However, these gains remain fragile and unable to deliver the sustained relief that the protracted crisis and their multidimensional complexities demand.^1,9^

Educational needs remain highly prevalent among Rohingya refugee children in Cox’s Bazar. Approximately a third of children age 3-18 were found to be in severe educational need and one in ten were experiencing extreme educational need. The findings reveal pronounced age-gender dynamics with teenage girls experiencing markedly higher educational needs, consistent with reported barriers related to deprioritisation, household survival pressures, sociocultural constraints and serious child protection concerns. Addressing the educational needs of Rohingya refugee children remains imperative to prevent gendered inequities, foster community resilience and strengthen recovery prospects from the protected crisis. Ameliorating the scale of children in educational need, their complexities and intersectoral dependencies requires strategic, intensive and sustained humanitarian relief efforts. Future longitudinal and qualitative research is needed to monitor educational needs, clarify causal pathways that influence individual severity scores and better capture sensitive protection-related barriers to education in consultation with community.

## Data Availability

The de-identified participant-level dataset underlying the study results are available at https://microdata.unhcr.org/index.php/catalog/1128.

## Acknowledgements

The authors wish to formally acknowledge those whose collective efforts have made this research article possible. Specifically, the Inter-Sector Coordination Group (ISCG), UNHCR, IOM, ECHO, the REACH Initiative, enumerators, other study personnel, and household respondents who graciously donated their time.

## Supporting information

**S1 Document**. STROBE checklist for cross-sectional studies.

## References

1. United Nations Office for the Coordination of Humanitarian Affairs (OCHA). Global Humanitarian Overview 2026: Rohingya Joint Response Plan (JRP). 2026 [cited 2026 Jan 8]. Available from: https://humanitarianaction.info/document/global-humanitarian-overview-2026/article/rohingya-joint-response-plan-jrp-1.

2. United Nations Human Rights Council. Situation of human rights of Rohingya Muslims and other minorities in Myanmar. 2025. Available from: https://digitallibrary.un.org/record/4084836.

3. Médecins Sans Frontières. The illusion of choice: Rohingya voices echo from the camps. 2025. Available from: https://reliefweb.int/report/bangladesh/illusion-choice-rohingya-voices-echo-camps.

4. Inter-Sector Coordination Group (ISCG). Bangladesh: Rohingya Refugee Response - Flash Appeal for 150,000 New Arrivals (June - December 2025). 2025. Available from: https://reliefweb.int/report/bangladesh/bangladesh-rohingya-refugee-response-flash-appeal-150000-new-arrivals-june-december-2025.

5. Inter-Sector Coordination Group (ISCG). Bangladesh: Rohingya Humanitarian Crisis Joint Response Plan January 2025 - December 2026. 2025. Available from: https://reliefweb.int/report/bangladesh/bangladesh-rohingya-humanitarian-crisis-joint-response-plan-january-2025-december-2026.

6. United Nations General Assembly. Situation of human rights of Rohingya Muslims and other minorities in Myanmar: report of the United Nations High Commissioner for Human Rights. 2025. Available from: https://digitallibrary.un.org/record/4092194/files/A_HRC_60_20-EN.pdf.

7. World Food Programme (WFP). Impact of Cuts - Bangladesh: Outcome of 2023 pilot study: Impact of funding shortfalls on beneficiaries. 2024. Available from: https://reliefweb.int/report/bangladesh/impact-cuts-bangladesh-outcome-2023-pilot-study-impact-funding-shortfalls-beneficiaries-august-2024.

8. The World Bank, World Food Programme (WFP). Impact of Food Ration Cuts on the Displaced Rohingya Population in Cox’s Bazar. 2024. Available from: https://reliefweb.int/report/bangladesh/bangladesh-impact-food-ration-cuts-displaced-rohingya-population-coxs-bazar-may-2024.

9. Inter-Sector Coordination Group (ISCG). Rohingya Refugee Response Urgent priorities: Addressing the Most Pressing Needs of the Joint Response Plan. 2025. Available from: https://reliefweb.int/report/bangladesh/bangladesh-rohingya-refugee-response-urgent-priorities-addressing-most-pressing-needs-joint-response-plan-june-december-2025.

10. REACH. Assessment of the Education Sector response to the Rohingya crisis - Rohingya refugee response. 2021. Available from: https://reliefweb.int/report/bangladesh/bangladesh-assessment-education-sector-response-rohingya-crisis-rohingya-refugee.

11. Cox’s Bazar Education Sector. Joint Education Needs Assessment: Rohingya Refugee in Cox’s Bazar - June 2018. 2018. Available from: https://reliefweb.int/report/bangladesh/joint-education-needs-assessment-rohingya-refugee-cox-s-bazar-june-2018.

12. Education Cannot Wait. Holistic learning outcomes measurement handbook for education in emergencies and protracted crises. 2025. Available from: https://reliefweb.int/report/world/holistic-learning-outcomes-measurement-handbook-education-emergencies-and-protracted-crises.

13. Inter-agency Network for Education in Emergencies (INEE). INEE Minimum Standards for Education: Preparedness, Response, Recovery (2024 Edition). 2024. Available from: https://reliefweb.int/report/world/inee-minimum-standards-education-preparedness-response-recovery-2024-edition.

14. Inter-agency Network for Education in Emergencies. Mind the Gap 3: Equity and Inclusion in and through Girls’ Education in Crisis. 2023. Available from: https://reliefweb.int/report/world/mind-gap-3-equity-and-inclusion-and-through-girls-education-crisis-enpt.

15. Østby G, Gjerløw H, Karim S, Dunlop E. Left Further Behind after the COVID-19 School Closures: Survey Evidence on Rohingya Refugees and Host Communities in Bangladesh. Journal on Education in Emergencies. 2023;9(1). doi: 10.33682/a1zn-5nda.

16. Wilson HJ. Drivers and determinants of extreme humanitarian needs among Rohingya refugee households: Evidence from UNHCR’s multi-sectoral needs analysis. PLOS ONE. 2025;20(12):e0331727. doi: 10.1371/journal.pone.0331727.

17. United Nations High Commissioner for Refugees (UNHCR), International Organization for Migration (IOM), Directorate-General for European Civil Protection and Humanitarian Aid Operations (ECHO). Bangladesh: Joint Multi Sector Needs Assessment: Cox’s Bazar, Rohingya Refugee Response – 2023, Refugees. 2023. Available from: https://microdata.unhcr.org/index.php/catalog/1128.

18. REACH. Bangladesh J-MSNA - MSNI Methodological Overview. 2023 [cited 2025 Apr 4]. Available from: https://repository.impact-initiatives.org/document/impact/474ac9e5/BGD2301_2023-J-MSNA_-MSNI-Methodological-overview_Jan-2024.pdf.

19. REACH. Research Terms of Reference: Joint Multi-Sector Needs Assessment MSNA 2023, Bangladesh. 2023 [cited 2025 Apr 4]. Available from: https://reliefweb.int/report/bangladesh/research-terms-reference-joint-multi-sector-needs-assessment-msna-2023-bgd2301-bangladesh-august-2023-v-1.

20. United Nations Office for the Coordination of Humanitarian Affairs (UNOCHA). Joint and Intersectoral Analysis Framework (JIAF) 2 Technical Manual. 2024. Available from: https://reliefweb.int/report/world/joint-and-intersectoral-analysis-framework-jiaf-2-technical-manual-july-2024.

21. Global Education Cluster. Calculating Education Cluster People in Need (PiN). 2024. Available from: https://www.educationcluster.net/calculating-people-need.

22. Inter-Sector Coordination Group (ISCG). Joint Multi-Sector Needs Assessment (J-MSNA): Bangladesh Rohingya Refugees - December 2021. 2022. Available from: https://reliefweb.int/report/bangladesh/joint-multi-sector-needs-assessment-j-msna-bangladesh-rohingya-refugees-december-2021.

23. World Food Programme (WFP). Bangladesh: Refugee Influx Emergency Vulnerability Assessment (REVA-8) Report, June 2025. 2025. Available from: https://reliefweb.int/report/bangladesh/bangladesh-refugee-influx-emergency-vulnerability-assessment-reva-8-report-june-2025.

24. Inter-Sector Coordination Group (ISCG). Joint Multi-Sector Needs Assessments (J-MSNA) Factsheet - Refugee Camp-level findings - December. 2023. Available from: https://reliefweb.int/report/bangladesh/bangladesh-joint-multi-sector-needs-assessment-j-msna-camp-level-findings-december-2023.

25. The Washington Group on Disability Statistics. The Washington Group Short Set on Functioning (WG-SS). 2022 [cited 2025 Apr 4]. Available from: https://www.washingtongroup-disability.com/question-sets/wg-short-set-on-functioning-wg-ss/.

26. StataCorp. Survey Data Reference Manual. 2025. Available from: https://www.stata.com/bookstore/survey-data-reference-manual/.

27. World Food Programme (WFP). Livelihood Coping Strategies Indicator for Food Security: List of strategies and their definitions. 2023 [cited 2025 June 6]. Available from: https://docs.wfp.org/api/documents/WFP-0000147820/download/.

28. Inter-Sector Coordination Group (ISCG). Bangladesh: Rohingya Humanitarian Crisis Joint Response Plan 2025-26 funding update as of 31 October 2025. 2025. Available from: https://reliefweb.int/report/bangladesh/bangladesh-rohingya-humanitarian-crisis-joint-response-plan-2025-26-funding-update-31-october-2025.

29. United Nations Children’s Fund (UNICEF). Humanitarian Action for Children 2026 - Bangladesh. 2026. Available from: http://reliefweb.int/report/bangladesh/humanitarian-action-children-2026-bangladesh.

